# Longitudinal White Matter Changes in Concussed Adolescents with Adverse Childhood Experiences

**DOI:** 10.64898/2026.07.22.26358355

**Authors:** Finian Keleher, Adrian I Onicas, Kevin C Bickart, Christine L Mac Donald, Anne Brown, Lawrence Cook, Frederick P Rivara, Gerard A Gioia, Christopher C Giza, Emily L Dennis, the Concussion Assessment, Research, and Education for Kids (CARE4Kids) consortium

## Abstract

Traumatic brain injury (TBI) is a leading cause of death and long-term disability in children, with many experiencing persistent symptoms even after mild TBIs. Exposure to adverse childhood experiences (ACEs) can have physiological effects that may alter how the brain responds to injury, yet the effects of ACEs on white matter injury and recovery processes remain unclear. This study examined whether a history of ACEs is associated with patterns of longitudinal change in white matter microstructure in children with mTBI. Ninety-six concussed adolescents (mean age=14.9 years, range =11.4–17.9, 51% female) from the CARE4Kids consortium completed the Pediatric ACEs and Related Life-Events Screener and underwent diffusion-weighted MRI at baseline (7-35 days after injury) and follow-up (2 months later). Fractional anisotropy (FA), mean diffusivity (MD), axial diffusivity (AD), radial diffusivity (RD), orientation dispersion index (ODI), and intracellular volume fraction (ICVF) were estimated using tract-based spatial statistics and harmonized across sites. Differences in the magnitude and direction of change in diffusion metrics over time were examined in 15 tracts of interest. Higher ACE exposure was associated with smaller absolute change in AD, MD, ODI, and ICVF across several white matter tracts, including the corpus callosum, internal and external capsules, corona radiata, and posterior thalamic radiation. Groups did not differ in the direction of white matter change for any tract-metric combination. These findings suggest that ACE exposure may blunt white matter reactivity to injury and/or reorganization during recovery processes.

## Introduction

Traumatic brain injury (TBI) is a significant public health issue for children and adolescents, with the prevalence in the United States estimated to be over 2 million.^1^ Although mild TBIs (mTBI) or concussions were traditionally considered to have primarily short term effects, they are increasingly recognized for their potential long-term consequences. Indeed, the field of TBI research is moving away from the terminology of “mild, moderate, and severe” precisely because it often does not accurately characterize the burden.^2^ Between 15% and 30% of children with “mTBIs” have been shown to develop long-lasting symptoms.^3^ However, our ability to predict who may develop persistent symptoms is poor. Additional research is needed to identify early predictors of long-term outcomes, psychosocial variables that may increase risk, and patterns of symptom progression over time. Understanding these factors could improve early identification of at-risk children, guide intervention strategies, and ultimately reduce the burden of pediatric mTBI.

One significant psychosocial risk factor that may influence development and recovery trajectories after TBI is childhood trauma exposure. Adverse childhood experiences (ACEs) can have behavioral consequences that may interact with TBI symptoms and complicate recovery, including emotional dysregulation, impaired stress responses, and disruptions in reward processing.^4,5^ Additionally, ACEs have been associated with diverse neurodevelopmental effects, such as volume reductions in subcortical structures, cortical thinning, and disruptions in functional connectivity.^6–8^ ACEs are also associated with alterations in biological systems central to injury responses, including alterations in neuroplasticity, chronic low-grade neuroinflammation, and blood-brain barrier disruption.^9–11^ Importantly, many of the systems affected by ACE exposure are primary mediators of injury and recovery processes following TBI.

Two primary theoretical frameworks exist to explain how the developmental effects of ACEs may influence the brain’s vulnerability to future environmental stressors. The cumulative risk or “double hit” model proposes that early life stress and trauma exposure compounds subsequent insults,^12^ leading to greater biopsychosocial disruption when a second stressor, such as mTBI, occurs. Alternatively, the calibration model suggests that early life adversity causes biological systems to adapt to harsh environments, effectively dampening biological responses to future stressors.^13–16^ Applied to TBI, these models generate competing predictions. Under a cumulative risk framework, ACE exposure would be expected to increase neural vulnerability, resulting in greater injury-related disruption. Under a calibration model framework, ACE-related adaptations in plasticity and inflammatory systems may instead blunt injury and recovery processes, leading to less overall change.

White matter microstructure provides a particularly relevant substrate for evaluating these competing models. Structural changes in the white matter of patients with mTBI are increasingly recognized for their contribution to the severity and persistence of symptoms.^17,18^ Compared to gross volumetric changes, microstructural alterations in the white matter may capture more subtle or diffuse effects associated with injury.^19^ The pathophysiological effects of ACEs on the brain suggest that ACE exposure and mTBI may interact to influence recovery of white matter microstructure following the injury.^16,20^ Most importantly, white matter degeneration and recovery after TBI are influenced by systems shown to be altered by ACE exposure, including neuroplasticity, neuroinflammation, and blood-brain barrier integrity.^9–11,21^ Children with a history of ACEs also often exhibit preexisting white matter abnormalities in tracts that are vulnerable to injury after TBI.^22–24^ Because ACE-related adaptations directly affect biological mechanisms mediating white matter injury and remodeling, the calibration model may provide a better framework for understanding the interactions between ACEs and mTBI. Rather than simply amplifying tissue damage, prior ACE-related adaptations may cause the brain to be less plastic, leading it to be simultaneously less reactive to injury and less capable of recovery processes.

The current study investigated whether exposure to ACEs is associated with patterns of structural brain changes during concussion recovery in a sample of children with mTBI from the CARE4Kids Study.^25^ The CARE4Kids study is a six-site study investigating multimodal predictors of persistent post-concussive symptoms, and includes diffusion weighted imaging scans collected within 5 weeks of injury and in the subacute phase of injury. We hypothesized that children with greater ACE exposure would exhibit a reduced magnitude of change in white matter microstructure over time, consistent with a calibration model, where systemic changes associated with ACE exposure render the brain less reactive to injury, but also less capable of recovery, after mTBI. If supported, these results would suggest that ACE exposure blunts, rather than amplifies, the brain’s response to mTBI. By investigating if early-life adversity influences the trajectory of structural brain alterations after TBI, this study has the potential to improve prognostic models and inform more targeted interventions for children that may be most at risk for prolonged symptoms.

## Methods

### Participants

Participants were enrolled as part of the Concussion Assessment, Research, and Education for Kids (CARE4Kids) consortium at five of the six sites that contributed diffusion-weighted imaging data to the present study (University of Washington/Seattle Children’s, University of California at Los Angeles, University of Texas Southwestern Medical Center, University of Rochester, and Atrium Health Wake Forest Baptist Hospital). Sites for recruitment included primary care clinics, emergency departments, concussion subspecialty clinics, athletic leagues, and schools. Eligible participants included English speakers ages 11–17.99 years diagnosed with a concussion by a health care provider using the Concussion in Sport Group criteria.^26^ Additionally, participants were included only if they continued to experience post-concussion symptoms at the time of first evaluation (T1) (7–35 days after injury). Participants with a history of developmental disorders, neurological disorders, severe psychiatric illness, substance abuse, moderate or severe TBI, or prior concussion (within the last 3 months) were excluded from the study. While the study had recruited 360 participants, only 96 adolescents were eligible for the current analysis since only a subset of patients underwent imaging and the ACE questions were only added partway through the cohort recruitment period.

The study was reviewed and approved by the University of Utah Institutional Review Board (IRB) under a single IRB. All participants provided written or verbal informed assent, while parents provided written informed consent.

### Procedure

The initial visit (T1) took place 7–35 days after injury. Participants underwent the complete CARE4Kids consortium test battery, which included neuropsychiatric assessments, symptom questionnaires, autonomic testing, and blood acquisition. Following the clinical assessment, participants completed an MRI neuroimaging protocol modeled after the Adolescent Brain Cognitive Development (ABCD) study, including T1-weighted and T2-weighted structural MRI, diffusion-weighted MRI, susceptibility-weighted imaging, resting-state functional MRI, and arterial spin labeling.^27^ Approximately three months later, participants completed a follow-up visit (T3) that included the identical clinical and neuroimaging protocols.

### Measures

Adverse childhood experience exposure was assessed using the Pediatric Adverse Childhood Experiences and Related Life-events Screener (PEARLS), a 17-item screening tool developed by Koita et al. (2018) for use in pediatric primary care settings to identify exposure to adverse childhood experiences and related life events.^28^ The PEARLS yields two summary scores: a total ACE score and a total life events score; individual item responses are not retained. The tool has demonstrated adequate to high internal consistency (Cronbach’s alpha = 0.81 and 0.82).^29,30^ In the present study, PEARLS data were collected via parent-proxy report at the initial visit.^28–30^

### Image Acquisition and Processing

Imaging was acquired across five sites on 3 Tesla scanners from three manufacturers: Siemens Prisma (University of California at Los Angeles, University of Rochester, University of Texas Southwestern Medical Center), Siemens Skyra (Atrium Health Wake Forest Baptist Hospital), and Philips Ingenia Elition (University of Washington/Seattle Children’s). To minimize cross-site variability, acquisition protocols were harmonized across sites and modeled after the Adolescent Brain Cognitive Development (ABCD) study.^27^ High-resolution T1-weighted structural images were acquired at 1.0 mm isotropic resolution. Diffusion-weighted images were acquired using a multi-shell protocol at 1.7 mm isotropic resolution, with diffusion weighting applied along 96 gradient directions distributed across four b-values (500, 1000, 2000, and 3000 s/mm²) and seven non-diffusion-weighted (b = 0) volumes. Site-specific acquisition parameters are presented in **Supplementary Table 1**. Raw imaging data from all five sites were transferred to the University of Utah for centralized processing and analysis.

Preprocessing was conducted at the University of Utah and included eddy-current correction and correction of echo-planar imaging-induced distortions. All data were visually checked for quality at multiple stages following ENIGMA-DTI and Neuroimaging Informatics Tools and Resources Clearinghouse recommendations, with particular attention to image registration. Two complementary models were then fit to the multi-shell data: the diffusion tensor model, which yielded fractional anisotropy (FA), mean diffusivity (MD), radial diffusivity (RD), and axial diffusivity (AD), and the Neurite Orientation Dispersion and Density Imaging (NODDI) model, which yielded orientation dispersion index (ODI) and intracellular volume fraction (ICVF). FA reflects the extent to which water diffusion is oriented along axonal pathways and is often used as an indirect marker of white matter organization and fiber coherence. MD measures the overall magnitude of diffusion within a voxel and can indicate cellularity or white matter damage. RD indexes diffusion perpendicular to the principal eigenvector, generally across the axon, and can reflect myelin integrity and white matter damage. AD captures diffusion parallel to the principal fiber direction and is often used to represent axonal integrity. ODI quantifies the angular spread of neurite orientations within a voxel, capturing microstructural restriction of water diffusion. ICVF estimates the proportion of restricted intracellular water, indicating axonal density.

The resulting metric maps were processed using tract-based spatial statistics (TBSS) through FSL software, in which maps were aligned to the ENIGMA-DTI template, projected onto the white-matter skeleton, and averaged within 15 regions of interest from the Johns Hopkins atlas.^31–33^ Some ROIs overlap (e.g., the genu [GCC], body [BCC], and splenium of the corpus callosum, as well as total corpus callosum, see enigma.ini.usc.edu/protocols/dti-protocols/).

### Statistical Analysis

Statistical analyses were conducted using RStudio v4.4.1. Data were harmonized using longitudinal ComBat, preserving the variability associated with age and sex. Absolute and signed change in FA, MD, RD, AD, ODI, and ICVF between baseline and follow-up were computed as the difference between follow-up and initial values.^34^ Participants whose difference score exceeded 3 standard deviations from the group mean were excluded from the corresponding tract-metric analysis. For the absolute change analysis, the absolute value of the difference score was then z-scored prior to analysis. For the signed change analysis, signed difference scores were z-scored.

PEARLS scores in the present sample were highly positively skewed, with 33.3% of participants reporting zero adverse childhood experiences and a median score of 1 (interquartile range: 0 to 3.25, range: 0 to 10), consistent with the floor-concentrated distributions commonly reported in pediatric samples.^35^ Given the non-linear distribution of scores, participants were classified into two groups using a threshold of 2 or more adverse childhood experiences, an approach that has been adopted in pediatric research when sample distributions are concentrated at low scores and higher thresholds yield markedly unbalanced groups.^36–38^ This threshold yielded a low-exposure group (score of 1 or zero; n = 55, 57.3%) and a high-exposure group (score of 2 or higher; n = 41, 42.7%), representing the most balanced group partition in this sample, as thresholds of 3 or more and 4 or more produced progressively more unequal distributions (33.3% and 25.0% in the high-exposure group, respectively). Standardized scores were then entered as outcomes in linear regression models using the base R lm function, with age at baseline and sex as covariates. FDR correction was applied to adjust for multiple comparisons across ROIs. To examine the robustness of findings to this analytical decision, complementary analyses were conducted using the continuous PEARLS score as a predictor.

## Results

### Participant Characteristics

This analysis included 96 participants, 47 male/49 female, mean age 14.9 years, age range=11.4-17.9 years. See **Table 1** for additional demographic and clinical characteristics.

**Table 1.** Participant demographics and clinical characteristics. Values are presented as mean (SD) or n (%). PEARLS = Pediatric ACEs and Related Life-events Screener. RAPID = Retrospective Adjusted Post-Injury Difference. T1 = baseline assessment (7–35 days post-injury); T2 = follow-up assessment.

| Variable | Full Sample (n=96) | ≤1 ACE (n=55) | ≥2 ACE (n=41) |
| --- | --- | --- | --- |
| **Continuous Variables, Mean ± SD [Range]** |  |  |  |
| Age at Baseline (years) | 14.9 ± 1.9 [11.4–17.9] | 14.7 ± 1.9 [11.5–17.8] | 15.2 ± 1.9 [11.4–17.9] |
| PEARLS Score at initial assessment | 2.2 ± 2.6 [0.0–10.0] | 0.4 ± 0.5 [0.0–1.0] | 4.5 ± 2.4 [2.0–10.0] |
| Time Since Injury (days) | 21.5 ± 8.7 [7.0–35.0] | 21.2 ± 9.0 [7.0–35.0] | 21.8 ± 8.4 [7.0–35.0] |
| RAPID Overall (T1) | 29.7 ± 21.4 [1.0–82.0] | 27.6 ± 21.5 [1.0–73.0] | 32.5 ± 21.2 [1.0–82.0] |
| RAPID Overall (T3) | 8.5 ± 15.0 [0.0–81.0] | 7.8 ± 15.6 [0.0–81.0] | 9.4 ± 14.3 [0.0–55.0] |
| **Biological Sex, n (%)** |  |  |  |
| Male | 47 (49.0) | 30 (54.5) | 17 (41.5) |
| Female | 49 (51.0) | 25 (45.5) | 24 (58.5) |
| **Race/Ethnicity, n (%)** |  |  |  |
| Asian | 3 (3.1) | 2 (3.6) | 1 (2.4) |
| Black or African American | 9 (9.4) | 6 (10.9) | 3 (7.3) |
| White, non-Hispanic | 67 (69.8) | 37 (67.3) | 30 (73.2) |
| More than one race | 10 (10.4) | 5 (9.1) | 5 (12.2) |
| Unknown or not reported | 7 (7.3) | 5 (9.1) | 2 (4.9) |
| **Mother's Education, n (%)** |  |  |  |
| < High school | 1 (1.0) | 0 (0.0) | 1 (2.4) |
| Some high school | 1 (1.0) | 1 (1.8) | 0 (0.0) |
| High school graduate or GED | 4 (4.2) | 1 (1.8) | 3 (7.3) |
| Vocational school or some college | 15 (15.6) | 11 (20.0) | 4 (9.8) |
| College degree | 27 (28.1) | 15 (27.3) | 12 (29.3) |
| Master's or doctoral degree | 19 (19.8) | 11 (20.0) | 8 (19.5) |
| No response | 29 (30.2) | 16 (29.1) | 13 (32.0) |
| **Mechanism of Injury, n (%)** |  |  |  |
| Sport/Recreation | 43 (44.8) | 27 (49.1) | 16 (39.0) |
| Fall (non-sport) | 8 (8.3) | 2 (3.6) | 6 (14.6) |
| Motor vehicle crash (MVC) | 7 (7.3) | 6 (10.9) | 1 (2.4) |
| Assaulted by someone | 2 (2.1) | 1 (1.8) | 1 (2.4) |
| Struck by/against object/person | 5 (5.2) | 2 (3.6) | 3 (7.3) |
| Other | 1 (1.0) | 0 (0.0) | 1 (2.4) |
| No response | 30 (31.3) | 16 (29.1) | 13 (31.7) |

ACE scores were positively skewed (median = 1, IQR = 0–3.25, range = 0–10), with n = 32 participants (33.3%) reporting no ACEs (**Figure 1, A**). Using a threshold of ≥2 ACEs classified 42.7% of the sample, indicating that this cutoff reflects moderate rather than high cumulative adversity in this cohort. The ≤ 1 ACE group had a median PEARLS score of 0 (IQR = 0–1), while the ACES≥2 group had a median PEARLS score of 4 (IQR = 3–5). The two groups did not differ on any diffusion metric across all 15 white matter tracts at baseline, before or after controlling for age and sex (all uncorrected *p* > 0.05; **Figure 1, B**).

**Figure 1.**
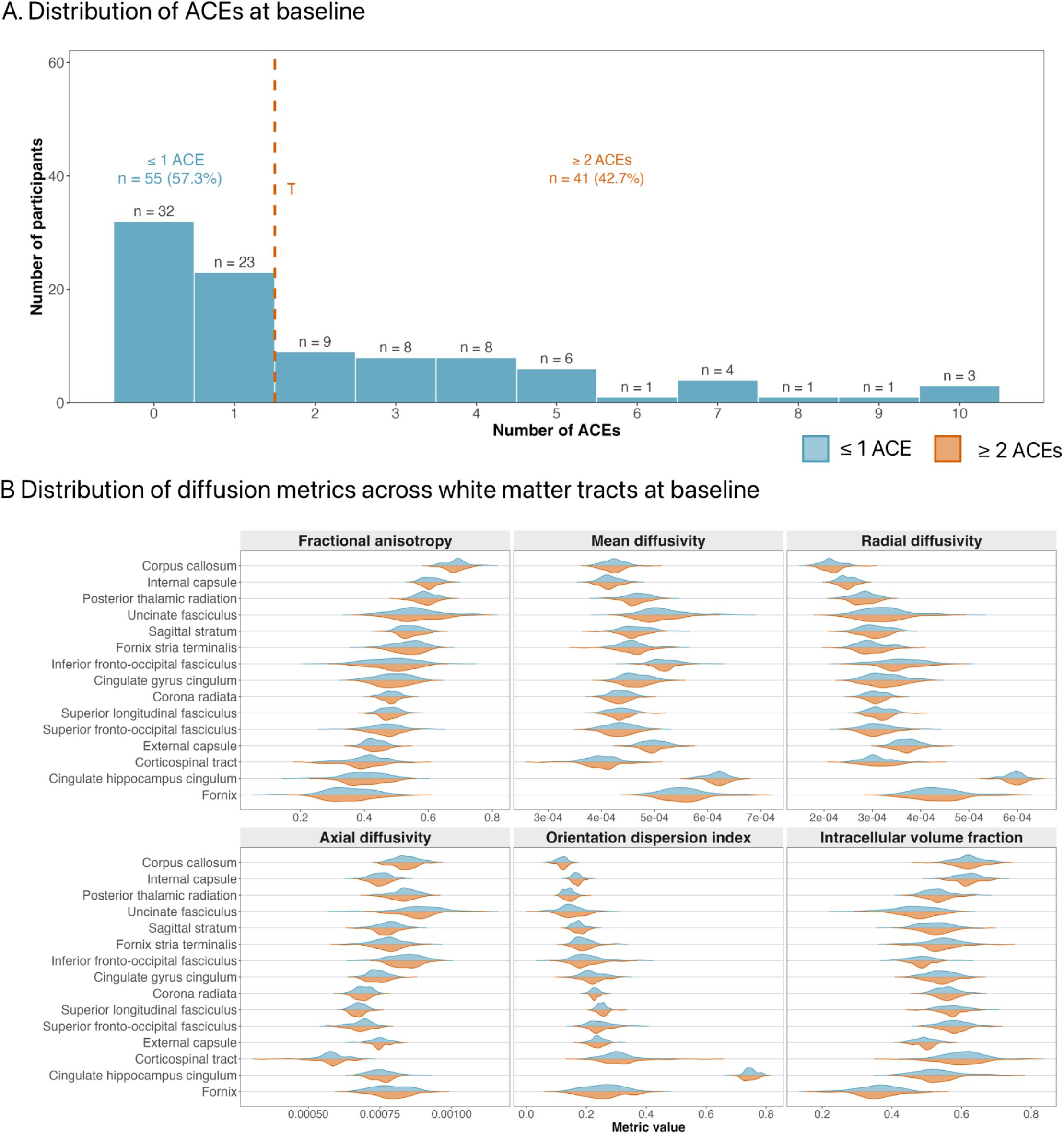
Distribution of PEARLS scores (A) and baseline diffusion metrics by ACE group (B). No between-group differences were observed for any tract across metrics at baseline, with or without controlling for age and sex (all *p* > 0.05 uncorrected).

### Absolute change

Significant differences between the ACE groups were found in the magnitude of change between T1 and T3 of AD, MD, ODI, and ICVF in several central white matter tracts, including the corpus callosum, external capsule, internal capsule, corona radiata, and posterior thalamic radiation. These results survived FDR correction for multiple comparisons (**Table 2, Figure 2**).

**Figure 2.**
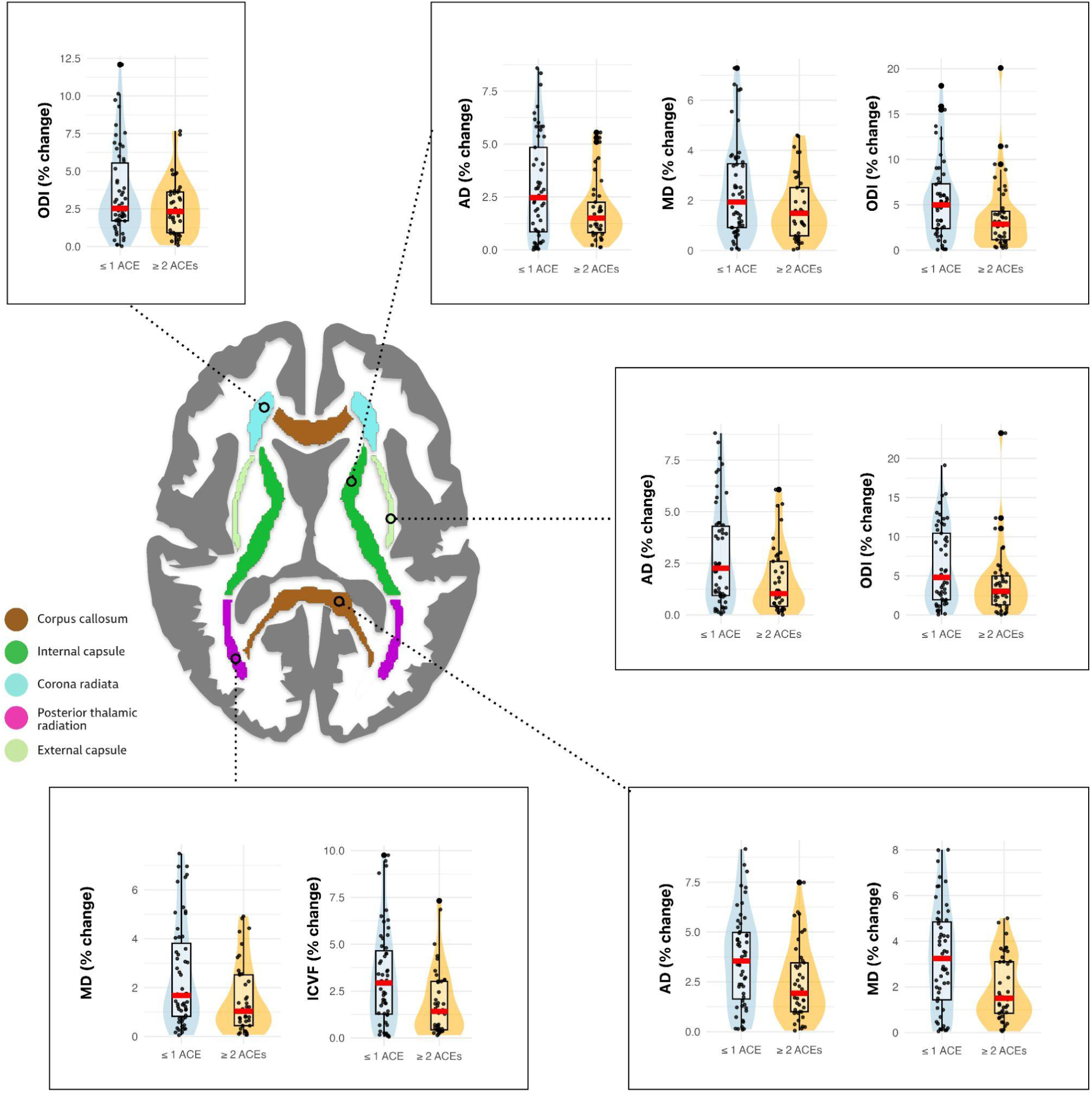
Associations between adverse childhood experiences (ACEs) and longitudinal changes in white matter microstructure following concussion. Violin plots show percentage change from baseline to follow-up in diffusion-weighted imaging metrics. Higher ACE exposure (≥ 2 ACEs) was associated with smaller absolute changes across time in axial diffusivity (AD), mean diffusivity (MD), orientation dispersion index (ODI), and intracellular volume fraction (ICVF) within the corpus callosum, internal capsule, corona radiata, posterior thalamic radiation, and external capsule.

**Table 2.** Group differences in absolute longitudinal changes. Beta values are standardized (z-scored absolute difference scores). Negative values indicate smaller absolute change in the high ACE exposure group (score of 2 or higher). FDR correction applied within each imaging modality (15 tests per modality).

| Metric | Tract | *beta* | *t* | *p* | *p*(FDR) |
| --- | --- | --- | --- | --- | --- |
| Axial diffusivity | Internal capsule | -0.626 | -3.10 | 0.003 | 0.039 |
|  | External capsule | -0.552 | -2.69 | 0.009 | 0.043 |
|  | Corpus callosum | -0.549 | -2.68 | 0.009 | 0.043 |
| Intracellular volume fraction | Posterior thalamic radiation | -0.633 | -3.12 | 0.002 | 0.036 |
| Mean diffusivity | Corpus callosum | -0.719 | -3.61 | <0.001 | 0.008 |
|  | Posterior thalamic radiation | -0.575 | -2.84 | 0.006 | 0.040 |
|  | Internal capsule | -0.554 | -2.71 | 0.008 | 0.040 |
| Orientation dispersion index | External capsule | -0.59 | -2.87 | 0.005 | 0.046 |
|  | Corona radiata | -0.563 | -2.81 | 0.006 | 0.046 |
|  | Internal capsule | -0.542 | -2.64 | 0.010 | 0.049 |

Axial diffusivity: Higher ACE exposure (ACEs ≥2) predicted smaller changes in AD of the corpus callosum (β = −0.549, t = −2.684, *p_FDR_* = 0.043), external capsule (β = −0.552, t = −2.690, *p_FDR_* = 0.043), and internal capsule (β = −0.626, t = −3.095, *p_FDR_*= 0.039).

Mean diffusivity: Higher ACE exposure (ACEs ≥2) was associated with smaller changes in MD of the corpus callosum (β = −0.719, t = −3.607, *p_FDR_* = 0.008), internal capsule (β = −0.554, t = −2.713, *p_FDR_* = 0.040), and posterior thalamic radiation (β = −0.575, t = −2.835, *p_FDR_*= 0.040).

Orientation dispersion index: For ODI, higher ACEs (ACEs ≥2) were associated with smaller changes in the corona radiata (β = −0.563, t = −2.808, *p_FDR_*= 0.046), external capsule (β = −0.590, t = −2.870, *p_FDR_* = 0.046), and internal capsule (β = −0.542, t = −2.643, *p_FDR_*= 0.048).

Intracellular volume fraction: Higher ACEs (ACEs ≥2) were associated with a lesser magnitude of change in ICVF in the posterior thalamic radiation (β = −0.633, t = −3.121, *p_FDR_*= 0.036).

Sensitivity analyses using the continuous PEARLS score as a predictor were consistent with the binary group results. Tract-metric associations identified in the primary analysis remained significant at the uncorrected level and in the same direction, the exception being mean diffusivity of the posterior thalamic radiation (p = 0.057; **Supplementary Table S2**).

### Signed change

No group differences in the direction of white matter change were observed (all *p* > 0.05 uncorrected).

## Discussion

The current study examined whether exposure to ACEs was associated with longitudinal changes in white matter microstructure after concussion in a sample of children with mTBI from the CARE4Kids Study. We hypothesized that children exposed to more ACEs would show lesser absolute change over time in DWI metrics than those exposed to one or no ACEs. We found less longitudinal change in AD, MD, ICVF, and ODI in several central white matter tracts in children who had experienced two or more ACEs in their lifetime. These results suggest that early-life adversity can indeed influence the trajectory of white matter alterations in adolescents with concussion, highlighting how psychosocial variables may contribute to recovery trajectories after mTBI. Further, these findings contribute to our understanding of how ACEs alter the brain’s response to future insult, providing leverage for evaluating two competing models that describe these interactions: the cumulative risk model and the calibration model. Specifically, the observed reductions in the magnitude of longitudinal white matter changes are more consistent with a calibration framework, in which systemic changes associated with ACEs render the brain less reactive to environmental stressors.

Notably, group differences emerged only in the magnitude, but not in the direction, of longitudinal changes in the white matter. This suggests that while children with higher ACE exposure exhibited less percent change in white matter metrics, the overall direction of those changes did not differ significantly between groups. This pattern does not support a cumulative risk model, which would predict amplified injury-related changes, and instead further supports a calibration model, in which the range of biological response is constrained. Accordingly, the present findings should be interpreted as differences in the magnitude of white matter reorganization rather than in its direction or outcome.

The diffusion metrics measured in the current study included FA, MD, RD, AD, ODI, and ICVF. No significant differences were found in FA or RD between ACE groups. However, significant differences were observed in the magnitude of absolute change in AD, MD, ODI, and ICVF metrics in several deep, central white matter tracts. We found less change in AD over time in the corpus callosum, internal capsule, and external capsule in children exposed to two or more ACEs.

AD measures the diffusion of water along axons and is generally interpreted to represent axonal structure and integrity. After TBI, AD can change as axons undergo progressive degeneration or reorganization. Mouse models of TBI have shown that AD is reduced in the acute phase of injury (<4 days), followed by a gradual return to more normative values in the 4 weeks following injury.^39^ However, in a study of mild TBI in humans, decreases in AD persisted 6 months after injury.^40^ These decreases in AD may reflect disruption or fragmentation of axonal structures, with later increases in AD values indicating restored axonal coherence. The lesser change in AD observed in the current study could therefore indicate less severe axonal degeneration during injury processes or disrupted axonal remodeling during recovery. However, given that T2 was approximately 2-3 months after injury, it is likely that differences in AD in the group with higher ACE exposure represent blunted post-traumatic microstructural response.

We also found less change in MD in the corpus callosum, external capsule, and posterior thalamic radiation among children exposed to ACEs. Because MD reflects the overall magnitude of water diffusion, it is influenced by multiple pathophysiological processes, making its interpretation more complex than that of AD or FA. Changes in MD are often inconsistent depending on the mechanism of and immunoresponse to injury. For example, MD may decrease in the presence of cytotoxic edema, which restricts diffusion as cellularity increases and axons become compressed, or increase in the presence of vasogenic edema, which increases extracellular volume.^41,42^ A lower change in MD could therefore reflect delayed resolution of edema, reduced white matter reorganization, or a blunted inflammatory response–depending on the phase of injury. Nonetheless, the majority of studies have found increases in MD after TBI, with these changes often persisting six months or more.^19,43,44^ Given that ACE exposure has been linked to chronic low-grade neuroinflammation and blood-brain barrier disruption, it is possible that children with high ACE exposure already exhibit increased MD at baseline.^10,11^ This effect has been observed previously, with ACE exposure and MD in several white matter tracts exhibiting a positive association.^45^ Consequently, the magnitude of MD change following injury may be less pronounced in this group compared to children who present with more normative inflammatory and recovery responses.

The magnitude of change in ODI was reduced in the corona radiata, internal capsule, and external capsule. ODI measures the angular spread of neurite orientations, indicating the overall coherence of fibers within a voxel. After TBI, ODI will generally increase, reflecting microstructural white matter disorganization. This has been demonstrated in patients with mTBI and moderate-severe TBI, with these changes often persisting into the chronic phase.^19,46^ During recovery processes, ODI will then decrease as axons realign and tract organization is restored.^47^ Lesser percent change in ODI may therefore represent reduced disruption in neurite orientation during injury processes or persistent disorganization and delayed recovery. While recovery timelines differ depending on mechanism and severity, a study of pediatric mTBI found that ODI returned to more normative levels 2-3 months after injury. Consequently, less absolute change in ODI among children with ACE exposure may reflect a restricted capacity to reorganize axonal orientation within the expected recovery window, consistent with the diminished plasticity known to occur after ACE exposure.^9,48^

Percent change in ICVF was also reduced in the corpus callosum and posterior thalamic radiation. ICVF is used to measure neurite density. After injury, white matter disruption may lead to less coherent and tightly packed axons, decreasing ICVF.^49^ Children with exposure to ACEs may have a lower baseline neurite density, such that they are less susceptible to axonal disruption but also have less capacity for structural recovery and remodeling. Chronic inflammation has also been linked to reduced neurite density and gliosis, which may further limit the degree of measurable change in ICVF following injury.^50^

All of the tracts with significant differences in absolute change in diffusion metrics between groups were deep, central white matter tracts. These central projection and commissural tracts facilitate long-range communication across cortical and subcortical networks and contribute to sensorimotor integration, cognitive control, emotional regulation, and interhemispheric transfer. They are also among regions commonly implicated in pediatric mTBI and in neurodevelopmental alterations related to ACEs exposure. The corpus callosum is particularly vulnerable to shear forces during TBI and has exhibited volume reductions and alterations in diffusion metrics following exposure to ACEs.^22,23,51^ Similarly, the internal and external capsules are known to be vulnerable to both TBI and microstructural damage associated with ACE exposure.^24,52,53^ The corona radiata is known to exhibit sustained changes in white matter microstructure after TBI, including reductions in FA and increases in RD.^23^ Lower FA has also been observed in the corona radiata in children exposed to childhood adversity.^54^ Lastly, the posterior thalamic radiation has been shown to have decreased structural organization following TBI and after childhood stress and trauma exposure.^52,55,56^ The similarities between ACE-related developmental alterations and TBI-related injury responses in these tracts suggest that pre-existing changes in white matter microstructure among children exposed to ACEs may limit the magnitude of additional microstructural change following TBI.

Across metrics measuring different components of white matter microstructure, a consistent pattern emerged in the current study: children with greater ACE exposure exhibited reduced longitudinal change in several central white matter tracts. Rather than amplifying tissue damage, as predicted by a cumulative risk model, these results suggest dampened injury and recovery processes, consistent with a calibration model.

Given the heterogeneity of the biological mechanisms that influence each diffusion metric, it is challenging to disentangle what the observed absolute differences reflect. Prior longitudinal studies indicate that microstructural abnormalities after pediatric mTBI can often persist for six months or more.^19,40,44^ Considering that follow-up in the present study occurred two to three months after injury, it is likely that the ACE-exposed group exhibited an attenuated white matter response to trauma. However, the absence of directional white matter changes limits the interpretability of individual DTI metrics, as the neurobiological meaning of metric-specific changes is contingent on whether values increased or decreased relative to baseline. Future work incorporating healthy controls, symptom assessments, and extended follow-up intervals will be essential for clarifying whether white matter injury or recovery processes are most affected by ACE exposure following adolescent concussion.

Although this study provides meaningful insight into the interactions between ACE exposure and mTBI, several limitations should be acknowledged. First, the current study did not include a non-injured comparison group, thus it does not isolate changes in white matter microstructure attributable to mTBI-related changes rather than other sources of variability. As longitudinal symptom data were not available, the observed white matter changes could not be assessed in relation to post-concussive symptoms. The present study included only a single follow-up timepoint, which limits the ability to characterize the temporal dynamics of white matter changes following mTBI in specific phases across time post-injury. Future studies incorporating multiple timepoints across the acute, subacute, and chronic phases of injury will be necessary to clarify recovery processes. Several limitations specific to the assessment of ACE exposure in the present sample are also worth mentioning. First, ACE scores in this sample were positively skewed, with few participants endorsing four or more ACEs, a threshold commonly associated with elevated health risk in the broader ACE literature. The relative absence of highly exposed individuals limits the ability to detect dose-response relationships and may reduce the generalizability of findings to populations with more severe cumulative adversity. Second, the PEARLS screener treats each endorsed item equivalently, such that different events receive equivalent weight, This approach does not account for differences in the type, severity, or chronicity of adverse experiences, and may obscure heterogeneity in how specific ACEs influence neurodevelopment and injury response. Third, ACE data in the present study were collected via parent-proxy report. Parents may not accurately report all aspects of their child’s adverse experiences, particularly when the parent is the source of the adversity.^57,58^ Future studies should consider supplementing parent report with child report to improve the completeness and validity of ACE assessment.

These findings suggest that early-life adversity shapes the brain’s response to concussion in ways that are more consistent with a calibration model than a cumulative risk model. Children with greater ACE exposure exhibited reduced longitudinal change in white matter microstructure across multiple metrics and central tracts, a pattern that reflects dampened rather than amplified injury and recovery processes. These results have implications for our understanding of how psychosocial history interacts with neurobiological responses to acute injury, and underscore the importance of considering early-life adversity when developing prognostic models for pediatric concussion. Future work incorporating healthy comparison groups, symptom outcomes, and extended longitudinal follow-up are needed to understand the consequences of this blunted microstructural response, specifically whether the lack of change confers protection against injury effects or reflects a limited capacity for recovery and higher risk of persistent symptoms. Ultimately, these findings add to a growing body of evidence suggesting that the brain’s response to concussion cannot be understood in isolation from its developmental history.

## Data Availability

All data produced in the present study are available upon reasonable request to the authors

## Acknowledgements

Funding for this study was provided by U54NS121688, R01NS122184, and R33NS120249. This work was conducted on behalf of the Concussion Assessment, Research, and Education for Kids (CARE4Kids) consortium. We thank all members for their contributions and support. The authors would like to thank the adolescents and families who participated in the CARE4Kids study.

## Supplementary Materials

**Supplementary Table S1.**
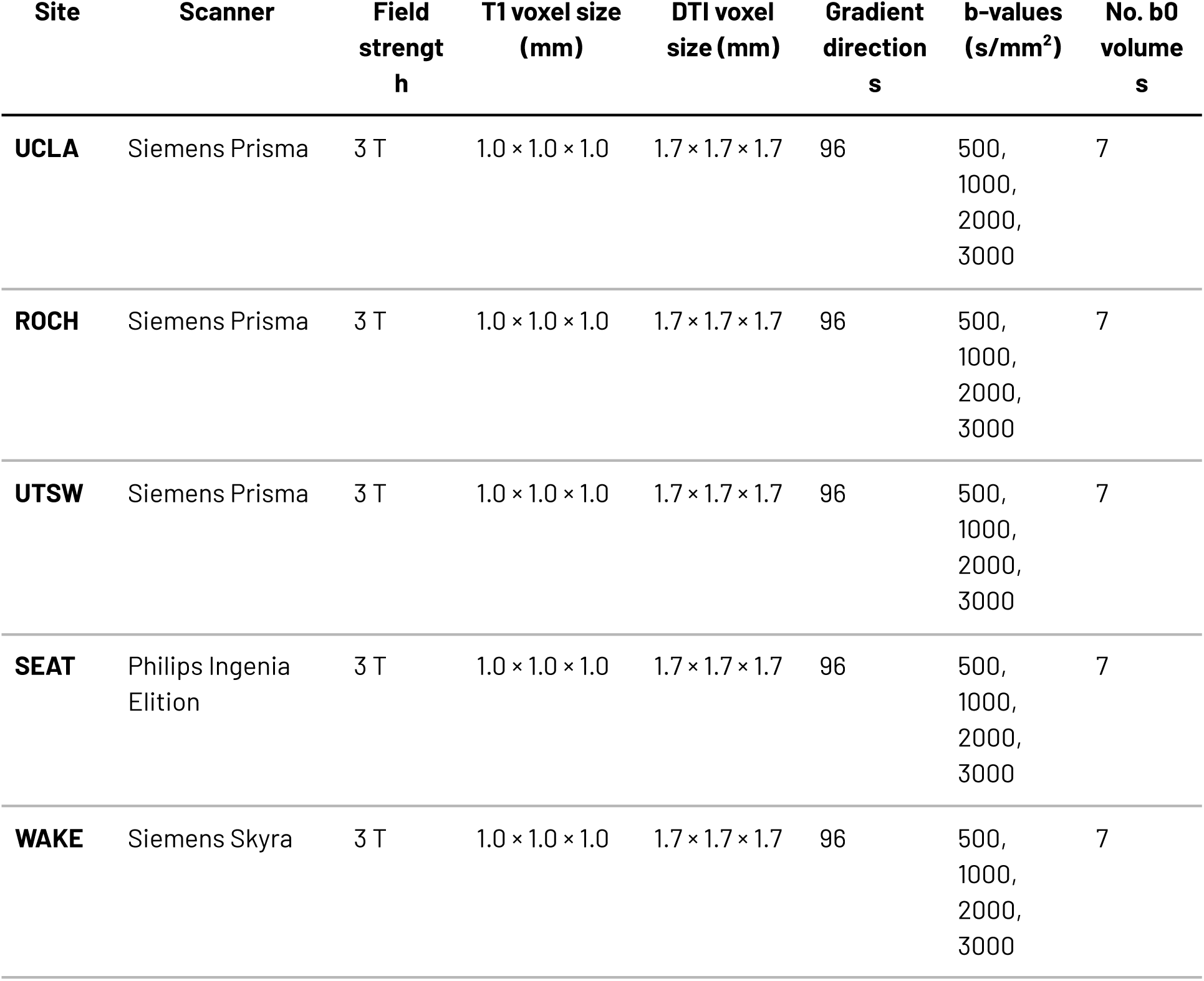
MRI Acquisition Parameters. Scan parameters for the five sites that contributed diffusion-weighted imaging data. UCLA = University of California, Los Angeles. ROCH = University of Rochester. UTSW = University of Texas Southwestern Medical Center. SEAT = University of Washington / Seattle Children’s Hospital. WAKE = Atrium Health Wake Forest Baptist Hospital.

**Supplementary Table S2.** Sensitivity analysis using continuous PEARLS score as predictor. Beta values are standardized (z-scored absolute difference scores).

| Metric | Tract | Beta | t | p |
| --- | --- | --- | --- | --- |
| Axial diffusivity | Internal capsule | -0.110 | -2.80 | 0.006 |
|  | Corpus callosum | -0.092 | -2.34 | 0.022 |
|  | External capsule | -0.081 | -2.02 | 0.046 |
| Intracellular volume fraction | Posterior thalamic radiation | -0.117 | -3.02 | 0.003 |
| Mean diffusivity | Corpus callosum | -0.109 | -2.77 | 0.007 |
|  | Internal capsule | -0.084 | -2.11 | 0.037 |
|  | Posterior thalamic radiation | -0.077 | -1.93 | 0.057 |
| Orientation dispersion index | Corona radiata | -0.087 | -2.23 | 0.028 |
|  | External capsule | -0.088 | -2.19 | 0.031 |
|  | Internal capsule | -0.085 | -2.12 | 0.037 |

## References

1 Lumba-Brown A, Yeates KO, Sarmiento K, Breiding MJ, Haegerich TM, Gioia GA, et al. Centers for Disease Control and Prevention Guideline on the Diagnosis and Management of Mild Traumatic Brain Injury Among Children. JAMA Pediatr 2018;172:e182853.

2 Manley GT, Dams-O’Connor K, Alosco ML, Awwad HO, Bazarian JJ, Bragge P, et al. A new characterisation of acute traumatic brain injury: the NIH-NINDS TBI Classification and Nomenclature Initiative. Lancet Neurol 2025;24:512–23.

3 Sheldrake E, Al-Hakeem H, Lam B, Goldstein BI, Wheeler AL, Burke M, et al. Mental health outcomes across the lifespan in individuals with persistent post-concussion symptoms: A scoping review. Front Neurol 2022;13:850590.

4 Tzouvara V, Kupdere P, Wilson K, Matthews L, Simpson A, Foye U. Adverse childhood experiences, mental health, and social functioning: A scoping review of the literature. Child Abuse Negl 2023;139:106092.

5 Landa-Blanco M, Vásquez G, Portillo G, Sproviero F, Echenique Y. The impact of adverse childhood experiences on mental health, sexual risk behaviors, and alcohol consumption in adulthood. Front Psychiatry 2024;15:1352824.

6 Schwartz A, Macalli M, Navarro MC, Jean FAM, Crivello F, Galera C, et al. Adverse childhood experiences and left hippocampal volumetric reductions: A structural magnetic resonance imaging study. J Psychiatr Res 2024;180:183–9.

7 Gold AL, Sheridan MA, Peverill M, Busso DS, Lambert HK, Alves S, et al. Childhood abuse and reduced cortical thickness in brain regions involved in emotional processing. J Child Psychol Psychiatry 2016;57:1154–64.

8 Holz NE, Berhe O, Sacu S, Schwarz E, Tesarz J, Heim CM, et al. Early social adversity, altered brain functional connectivity, and mental health. Biol Psychiatry 2023;93:430–41.

9 Smith KE, Pollak SD. Rethinking concepts and categories for understanding the neurodevelopmental effects of childhood adversity. Perspect Psychol Sci 2021;16:67–93.

10 Andersen SL. Neuroinflammation, early-life adversity, and brain development. Harv Rev Psychiatry 2022;30:24–39.

11 Falcone T, Janigro D, Lovell R, Simon B, Brown CA, Herrera M, et al. S100B blood levels and childhood trauma in adolescent inpatients. J Psychiatr Res 2015;62:14–22.

12 Kuhlman KR. Pitfalls and potential: Translating the two-hit model of early life stress from pre-clinical non-human experiments to human samples. Brain Behav Immun Health 2024;35:100711.

13 Evans GW, Li D, Whipple SS. Cumulative risk and child development. Psychol Bull 2013;139:1342–96.

14 Tooley UA, Bassett DS, Mackey AP. Environmental influences on the pace of brain development. Nat Rev Neurosci 2021;22:372–84.

15 Miller JG, Dennis EL, Heft-Neal S, Jo B, Gotlib IH. Fine particulate air pollution, early life stress, and their interactive effects on adolescent structural brain development: A longitudinal tensor-based morphometry study. Cereb Cortex 2022;32:2156–69.

16 McLaughlin KA, Weissman D, Bitrán D. Childhood adversity and neural development: A systematic review. Annu Rev Dev Psychol 2019;1:277–312.

17 McInnes K, Friesen CL, MacKenzie DE, Westwood DA, Boe SG. Mild Traumatic Brain Injury (mTBI) and chronic cognitive impairment: A scoping review. PLoS One 2017;12:e0174847.

18 Papini MG, Avila AN, Fitzgerald M, Hellewell SC. Evidence for altered white matter organization after mild traumatic brain injury: A scoping review on the use of diffusion magnetic resonance imaging and blood-based biomarkers to investigate acute pathology and relationship to persistent post-concussion symptoms. J Neurotrauma 2025;42:640–67.

19 Bourke NJ, Yanez Lopez M, Jenkins PO, De Simoni S, Cole JH, Lally P, et al. Traumatic brain injury: a comparison of diffusion and volumetric magnetic resonance imaging measures. Brain Commun 2021;3:fcab006.

20 Zhu J, MacIsaac-Jones MAC, Jenkins S, Yeates KO, Madigan S. Association between adverse childhood experiences score and traumatic brain injury occurrence: A systematic review and meta-analysis. J Neurotrauma 2025;42:1907–17.

21 Tianyi L, Huibregtse ME, Ely TD, van Rooij SJH, Lebois LAM, Webb EK, et al. Childhood adversity is associated with longitudinal white matter changes after adulthood trauma. medRxiv 2025. 10.1101/2025.03.08.25323425.

22 McCarthy-Jones S, Oestreich LKL, Lyall AE, Kikinis Z, Newell DT, Savadjiev P, et al. Childhood adversity associated with white matter alteration in the corpus callosum, corona radiata, and uncinate fasciculus of psychiatrically healthy adults. Brain Imaging Behav 2018;12:449–58.

23 Dinkel J, Drier A, Khalilzadeh O, Perlbarg V, Czernecki V, Gupta R, et al. Long-term white matter changes after severe traumatic brain injury: a 5-year prospective cohort. AJNR Am J Neuroradiol 2014;35:23–9.

24 Wong SA, Lebois LAM, Ely TD, van Rooij SJH, Bruce SE, Murty VP, et al. Internal capsule microstructure mediates the relationship between childhood maltreatment and PTSD following adulthood trauma exposure. Mol Psychiatry 2023;28:5140–9.

25 Giza CC, Gioia G, Cook LJ, Asarnow R, Snyder A, Babikian T, et al. CARE4Kids study: Endophenotypes of persistent post-concussive symptoms in adolescents: Study rationale and protocol. J Neurotrauma 2024;41:171–85.

26 Echemendia RJ, Meeuwisse W, McCrory P, Davis GA, Putukian M, Leddy J, et al. The Sport Concussion Assessment Tool 5th Edition (SCAT5): Background and rationale. Br J Sports Med 2017;51:848–50.

27 Casey BJ, Cannonier T, Conley MI, Cohen AO, Barch DM, Heitzeg MM, et al. The Adolescent Brain Cognitive Development (ABCD) study: Imaging acquisition across 21 sites. Dev Cogn Neurosci 2018;32:43–54.

28 Koita K, Long D, Hessler D, Benson M, Daley K, Bucci M, et al. Development and implementation of a pediatric adverse childhood experiences (ACEs) and other determinants of health questionnaire in the pediatric medical home: A pilot study. PLoS One 2018;13:e0208088.

29 Thakur N, Hessler D, Koita K, Ye M, Benson M, Gilgoff R, et al. Pediatrics adverse childhood experiences and related life events screener (PEARLS) and health in a safety-net practice. Child Abuse Negl 2020;108:104685.

30 Ye M, Hessler D, Ford D, Benson M, Koita K, Bucci M, et al. Pediatric ACEs and related life event screener (PEARLS) latent domains and child health in a safety-net primary care practice. BMC Pediatr 2023;23:367.

31 Smith SM, Jenkinson M, Johansen-Berg H, Rueckert D, Nichols TE, Mackay CE, et al. Tract-based spatial statistics: voxelwise analysis of multi-subject diffusion data. Neuroimage 2006;31:1487–505.

32 Jenkinson M, Beckmann CF, Behrens TEJ, Woolrich MW, Smith SM. FSL. Neuroimage 2012;62:782–90.

33 Mori S, Oishi K, Faria AV. White matter atlases based on diffusion tensor imaging. Curr Opin Neurol 2009;22:362–9.

34 Beer JC, Tustison NJ, Cook PA, Davatzikos C, Sheline YI, Shinohara RT, et al. Longitudinal ComBat: A method for harmonizing longitudinal multi-scanner imaging data. Neuroimage 2020;220:117129.

35 Crouch E, Probst JC, Radcliff E, Bennett KJ, McKinney SH. Prevalence of adverse childhood experiences (ACEs) among US children. Child Abuse Negl 2019;92:209–18.

36 Marie-Mitchell A, Watkins HBR, Copado IA, Distelberg B. Use of the Whole Child Assessment to identify children at risk of poor outcomes. Child Abuse Negl 2020;104:104489.

37 Patel NS, Watkins H, Marie-Mitchell A. Predictive validity of the Whole Child Assessment in a generally healthy pediatric cohort. J Prim Care Community Health 2023;14:21501319231168342.

38 Rodenbough A, Opolka C, Wang T, Gillespie S, Ververis M, Fitzpatrick AM, et al. Adverse childhood experiences and patient-reported outcome measures in critically ill children. Front Pediatr 2022;10:923118.

39 Mac Donald CL, Dikranian K, Bayly P, Holtzman D, Brody D. Diffusion tensor imaging reliably detects experimental traumatic axonal injury and indicates approximate time of injury. J Neurosci 2007;27:11869–76.

40 Palacios EM, Yuh EL, Mac Donald CL, Bourla I, Wren-Jarvis J, Sun X, et al. Diffusion tensor imaging reveals elevated diffusivity of white matter microstructure that is independently associated with long-term outcome after mild Traumatic Brain Injury: A TRACK-TBI study. J Neurotrauma 2022;39:1318–28.

41 Shenton ME, Hamoda HM, Schneiderman JS, Bouix S, Pasternak O, Rathi Y, et al. A review of magnetic resonance imaging and diffusion tensor imaging findings in mild traumatic brain injury. Brain Imaging Behav 2012;6:137–92.

42 Grossman EJ, Inglese M, Bammer R. Mild traumatic brain injury: is diffusion imaging ready for primetime in forensic medicine? Top Magn Reson Imaging 2010;21:379–86.

43 Khong E, Odenwald N, Hashim E, Cusimano MD. Diffusion tensor imaging findings in post-concussion syndrome patients after mild traumatic brain injury: A systematic review. Front Neurol 2016;7:156.

44 Veeramuthu V, Narayanan V, Kuo TL, Delano-Wood L, Chinna K, Bondi MW, et al. Diffusion tensor imaging parameters in mild traumatic brain injury and its correlation with early neuropsychological impairment: A longitudinal study. J Neurotrauma 2015;32:1497–509.

45 Poletti S, Mazza E, Bollettini I, Locatelli C, Cavallaro R, Smeraldi E, et al. Adverse childhood experiences influence white matter microstructure in patients with schizophrenia. Psychiatry Res 2015;234:35–43.

46 Cao M, Luo Y, Wu Z, Wu K, Li X. Abnormal neurite density and orientation dispersion in frontal lobe link to elevated hyperactive/impulsive behaviours in young adults with traumatic brain injury. Brain Commun 2022;4:fcac011.

47 Stein A, Vinh To X, Nasrallah FA, Barlow KM. Evidence of ongoing cerebral microstructural reorganization in children with persisting symptoms following mild traumatic brain injury: A NODDI DTI analysis. J Neurotrauma 2024;41:41–58.

48 Cross D, Fani N, Powers A, Bradley B. Neurobiological development in the context of childhood trauma. Clin Psychol (New York) 2017;24:111–24.

49 Wu Y-C, Mustafi SM, Harezlak J, Kodiweera C, Flashman LA, McAllister TW. Hybrid diffusion imaging in mild traumatic brain injury. J Neurotrauma 2018;35:2377–90.

50 Zhang W, Xiao D, Mao Q, Xia H. Role of neuroinflammation in neurodegeneration development. Signal Transduct Target Ther 2023;8:267.

51 Kim E, Seo HG, Lee HH, Lee SH, Choi SH, Cho W-S, et al. Altered white matter integrity after mild to moderate traumatic brain injury. J Clin Med 2019;8:1318.

52 Caeyenberghs K, Leemans A, Geurts M, Taymans T, Linden CV, Smits-Engelsman BCM, et al. Brain-behavior relationships in young traumatic brain injury patients: DTI metrics are highly correlated with postural control. Hum Brain Mapp 2010;31:992–1002.

53 Sibilia F, Custer RM, Irimia A, Sepehrband F, Toga AW, Cabeen RP, et al. Life after mild Traumatic Brain Injury: Widespread structural brain changes associated with psychological distress revealed with multimodal magnetic resonance imaging. Biol Psychiatry Glob Open Sci 2023;3:374–85.

54 Mitchell O, Roddy DW, Connaughton M. Early life adversity and white matter microstructural organization-a systematic review. Brain Imaging Behav 2025;19:785–99.

55 McManus E, Haroon H, Duncan NW, Elliott R, Muhlert N. The effects of stress across the lifespan on the brain, cognition and mental health: A UK biobank study. Neurobiol Stress 2022;18:100447.

56 Rodriguez A, Petropoulos H, Sanjuan PM, Wang Y-P, Wilson TW, Calhoun VD, et al. Childhood adversity and white matter microstructure: White matter differences associated with trauma exposure. Stresses 2025;5:19.

57 Kataja E-L, Tuulari JJ, Karlsson L, Sinisalo S, Autere T-A, Perasto L, et al. Measuring adverse childhood experiences by interviewing children at 9 and 10 years of age: Prevalence, concordance with mother-reports, posttraumatic stress disorder symptoms, and subjective experience of being asked about adverse childhood experiences in FinnBrain Birth cohort study. J Exp Child Psychol 2025;256:106238.

58 Caqueo-Urízar A, Urzúa A, Villalonga-Olives E, Atencio-Quevedo D, Irarrázaval M, Flores J, et al. Children’s mental health: Discrepancy between child self-reporting and parental reporting. Behav Sci (Basel) 2022;12:401.

